# U-shaped effect of blood pressure on structural OCT metrics and retinal blood flow autoregulation in ophthalmologically healthy subjects

**DOI:** 10.1101/2021.01.14.21249808

**Authors:** Konstantinos Pappelis, Nomdo M. Jansonius

**Author notes:** <u>Correspondence:</u> K. Pappelis, Dept. of Ophthalmology, University Medical Center Groningen, P.O.Box 30.001, 9700 RB Groningen, Netherlands. <u>E-mail co-authors:</u>. <u>Financial support:</u> European Union’s Horizon 2020 Innovative Training Networks Program, under the Marie Sklodowska – Curie grant, Project ID 675033. The funding organization had no role in the design, conduct, analysis, or publication of this research. <u>Meeting presentation:</u> European Association for Vision and Eye Research (EVER); Nice, 3rd-6th October 2018.

## Abstract

**Objective:** 1) To investigate the effect of low blood pressure (BP), treated arterial hypertension (AHT), and untreated AHT on the ganglion cell-inner plexiform layer (GCIPL) and the retinal nerve fiber layer (RNFL) thickness of non-glaucomatous eyes and 2) to elucidate whether this effect is related to crossing the lower limit of retinal blood flow (RBF) autoregulation.

**Design:** Cross-sectional, case-control.

**Subjects:** We included 96 eyes of 96 ophthalmologically healthy subjects (age 50-65). Participants were prospectively recruited from a large-scale cohort study in the northern Netherlands (n=167,000; Lifelines Biobank). They were allocated to four groups (low BP, normal BP [controls], treated AHT, untreated AHT), based on information from previous visits and strict distribution criteria.

**Methods:** Inner retinal layer thicknesses were obtained with optical coherence tomography (OCT). Fractal dimension of the superficial microvasculature was quantified with OCT-angiography and customized software. Central retinal vessel diameters were obtained from fundus images. BP and intraocular pressure measurements were also acquired. Measurements were combined with a validated physiological model to estimate vascular outcome measures. Structural and vascular metrics were compared across groups and mediation analysis was performed.

**Main outcome measures:** Structural: macular GCIPL and RNFL (mRNFL), peripapillary RNFL (pRNFL) thickness. Vascular: RBF, retinal vascular resistance (RVR), autoregulatory reserve (AR).

**Results:** Compared to controls, GCIPL was thinner in the low BP group (*P*=0.013), treated hypertensives (*P*=0.007), and untreated hypertensives (*P*=0.007). Treated hypertensives exhibited the thinnest mRNFL (*P*=0.001), temporal pRNFL (*P*=0.045), and inferior pRNFL (*P*=0.034). In multivariable analysis, RBF was mediating the association of GCIPL thickness with BP within the combined low BP group and controls (*P*=0.003), RVR together with AR were mediating the same association within the combined treated hypertensives and controls (*P*=0.001 and *P*=0.032), and RVR was mediating the association within the combined untreated antihypertensives and controls (*P*=0.022).

**Conclusions:** We uncovered GCIPL and RNFL thinning related to both tails of the BP distribution. GCIPL thinning was associated with reduced RBF autoregulatory capacity. This predisposition to glaucomatous damage could explain the frequent epidemiological finding of increased glaucoma risk in certain subgroups, such as subjects with nocturnal BP dipping or aggressively treated AHT. Longitudinal studies could confirm this postulation.

## Introduction

Glaucoma is a chronic optic neuropathy characterized by thinning of the retinal nerve fiber layer (RNFL), loss of retinal ganglion cells (RGCs), and progressive visual function decline.^1^ While elevated intraocular pressure (IOP) is considered as the most important modifiable risk factor, glaucoma may also manifest in those with apparently normal IOP (normal-tension glaucoma [NTG]).^2–4^ Therefore, certain components of the disease remain elusive or insufficiently addressed.

It has been proposed that low or unstable blood supply could lead to reduced oxygenation of the RGCs.^4,5^ Current assessment of RGC structure is based on Optical Coherence Tomography (OCT), while its extensions, OCT-angiography (OCT-A) and Doppler OCT, enable the non-invasive evaluation of retinal perfusion.^6–8^ These methods have already revealed that reduced blood flow predicts visual field (VF) deterioration, independently of neural tissue damage.^9–11^ However, after the onset of the disease, it is impossible to disentangle if perfusion deficits are the cause (low supply) or consequence (low demand) of glaucomatous optic neuropathy (GON). This realization is known as the ‘chicken-egg’ dilemma in glaucoma.

From a hemodynamic perspective, blood flow is determined by the balance between ocular perfusion pressure (OPP) and vascular resistance.^12^ Therefore, low blood pressure (BP) could result in low OPP, thus increasing the risk for glaucoma incidence and progression, possibly due to flow-mediated damage to the RGCs. Indeed, this has been observed in some cross-sectional and longitudinal population-based studies.^13–16^ However, other studies do not confirm this finding, while there is also evidence that this association becomes relevant only when low BP manifests as pronounced nocturnal dipping.^17,18^ On the other hand, while arterial hypertension (AHT) is also frequently reported as a risk factor for glaucoma, conflicting results exist on whether BP reduction exacerbates or protects from GON, possibly depending on individual medication effects and on how aggressive the treatment strategy is.^19–26^

This study focuses on ophthalmologically healthy subjects, rather than glaucoma subjects. This is essential, because the next logical step in approaching the ‘chicken-egg’ dilemma is to move back in the disease time course and study the effect of BP on retinal blood flow (RBF) and the RGCs, prior to any glaucoma onset. To investigate RBF, we look into the retinal microcirculation, which demonstrates the ability of autoregulation, i.e., active modification of vascular caliber in response to local signals.^27^ This property protects the tissue from ischemia, in case of OPP drops. Recently, we proposed and validated a microcirculation model predicting which healthy subjects are more prone to hypoperfusion, by estimating their lower autoregulation limit (LARL) through OCT-A, fundus imaging, and other clinical examinations.^28^

In the present study, we hypothesized an inverse U-shaped association between BP status and structural OCT measures in non-glaucomatous eyes. While the detrimental effect of AHT to the RGCs and their axons has been previously documented, this effect has not been studied in subjects with low BP, nor has it been examined in combination with RBF autoregulation.^29–32^ Studies until now have used linear models to describe the association between BP and structural OCT measures, thus potentially neglecting any signal coming from the left tail of the distribution.^33,34^

Therefore, the aims of this study were 1) to investigate the effect of low BP, treated AHT, and untreated AHT on the inner retinal layer thicknesses of non-glaucomatous eyes and 2) to elucidate whether this effect is related to crossing the lower limit of RBF autoregulation. For this purpose, we performed multimodal structural and vascular imaging in ophthalmically healthy normotensive controls, treated arterial hypertensives, and individuals belonging to the lower and higher (untreated) tails of the BP distribution. Participants were selected from the large-scale population-based Lifelines cohort, which enabled us to study the real extremes, especially from the low BP tail, in an unbiased manner.

## Methods

### Study design and population

For this cross-sectional, case-control study, we prospectively recruited subjects via targeted invitation among the participants of a large-scale prospective cohort study of the northern Netherlands (Lifelines Biobank; n=167,000).^35^ Subjects were invited based solely on their BP status and age. Following a strict selection procedure, 105 participants between 50 and 65 years of age satisfied both the BP criteria (see next paragraph) and the ophthalmic and medical history inclusion criteria: unoperated eyes; best-corrected visual acuity ≥ 0.8; spherical refractive error between -3 and +3 D; cylinder not exceeding 2 D; IOP ≤ 21 mmHg (non-contact tonometer Tonoref II, Nidek, Aichi, Japan); no reproducibly abnormal VF test locations (Frequency Doubling Technology [C20-1 screening mode], Carl Zeiss, Jena, Germany); no family history of glaucoma; no ophthalmic, hematologic, or cardiovascular disease (except for AHT), and no diabetes. We performed additional documentation of ophthalmic health with the subsequent imaging sessions (see *Data collection*). We allocated participants to four non-overlapping groups: 1) low BP, 2) normal BP (controls), 3) treated AHT, and 4) untreated AHT. Group definitions (see next paragraph) were based on previous epidemiological evidence on the association between BP status and glaucoma.^15,16^ All participants provided written informed consent. The ethics board of the University Medical Center Groningen approved the study protocol (#NL61508.042.17). The study followed the tenets of the Declaration of Helsinki.

### Blood pressure group definitions

We defined low BP (group 1) as both systolic and diastolic BP (SBP, DBP) lower than the 10th percentiles of the age-matched population (110 mmHg and 65 mmHg, respectively), without any AHT record. This criterion had to be confirmed on at least two previous, separate occasions (ascertaining that subjects truly belonged to the tail of the distribution and did not regress towards the mean). We defined untreated AHT (group 4) similarly, the criteria being both SBP and DBP higher than the 90th percentiles of the age-matched population (149 mmHg and 88 mmHg, respectively), verified at least twice previously. Subjects of this group were aware of their BP status, but never made use of antihypertensive medication, by choice. For treated AHT (group 3), we randomly invited participants documented as receiving (and still making uninterrupted use of) antihypertensive medication for at least one year. Lastly, we defined normal BP (group 2) as both SBP and DBP within 1 standard deviation (SD) from the mean of the age-matched population (SBP: 113 mmHg to 143 mmHg and DBP: 67 mmHg to 85 mmHg, measured on site) and no previous record of AHT.

### Data collection

All participants were examined at the same time of the day (5:00 PM-6:30 PM) and were not given any instructions regarding their routine prior to their visit. Following screening (see previous section), we applied mydriatic drops that have been shown to not affect RBF (tropicamide 0.5%).^36^ After the participants had rested in a quiet room for 20 minutes, we recorded BP from the brachial artery, in sitting position, with an automatic monitor (Omron M6 Comfort, Omron Healthcare, Kyoto, Japan). We averaged two readings, unless there was a discrepancy of at least 10 mmHg in SBP or 5 mmHg in DBP, in which case we averaged three readings. We also measured the weight and height of each participant.

For the imaging session, we selected, randomly if both eyes fulfilled the inclusion criteria, one eye per participant. We performed macular and optic nerve head (ONH) structural OCT imaging, as well as parafoveal OCT-A (Canon HS100 SD-OCT, Tokyo, Japan). The device automatically segments and quantifies macular RNFL (mRNFL) thickness and ganglion cell-inner plexiform layer (GCIPL) within a 10-mm diameter circular region of interest (ROI) centered at the fovea (Figure 1A), a region which has been shown to be advantageous over the commonly used 5-mm diameter ROI for mRNFL measurements.^37^ It also reports peripapillary RNFL (pRNFL) thickness at a 3.45-mm diameter circle centered at the ONH (Figure 1B). We further subdivided pRNFL into temporal, superior, nasal, and inferior. We additionally acquired two 6×6 mm OCT-A scans centered at the fovea (Figure 1C). We required an image quality of 7/10 or better, as well as the absence of any artifacts or segmentation errors for all OCT and OCT-A scans, resulting in the exclusion of 9 out of the 105 subjects.

**Figure 1.**
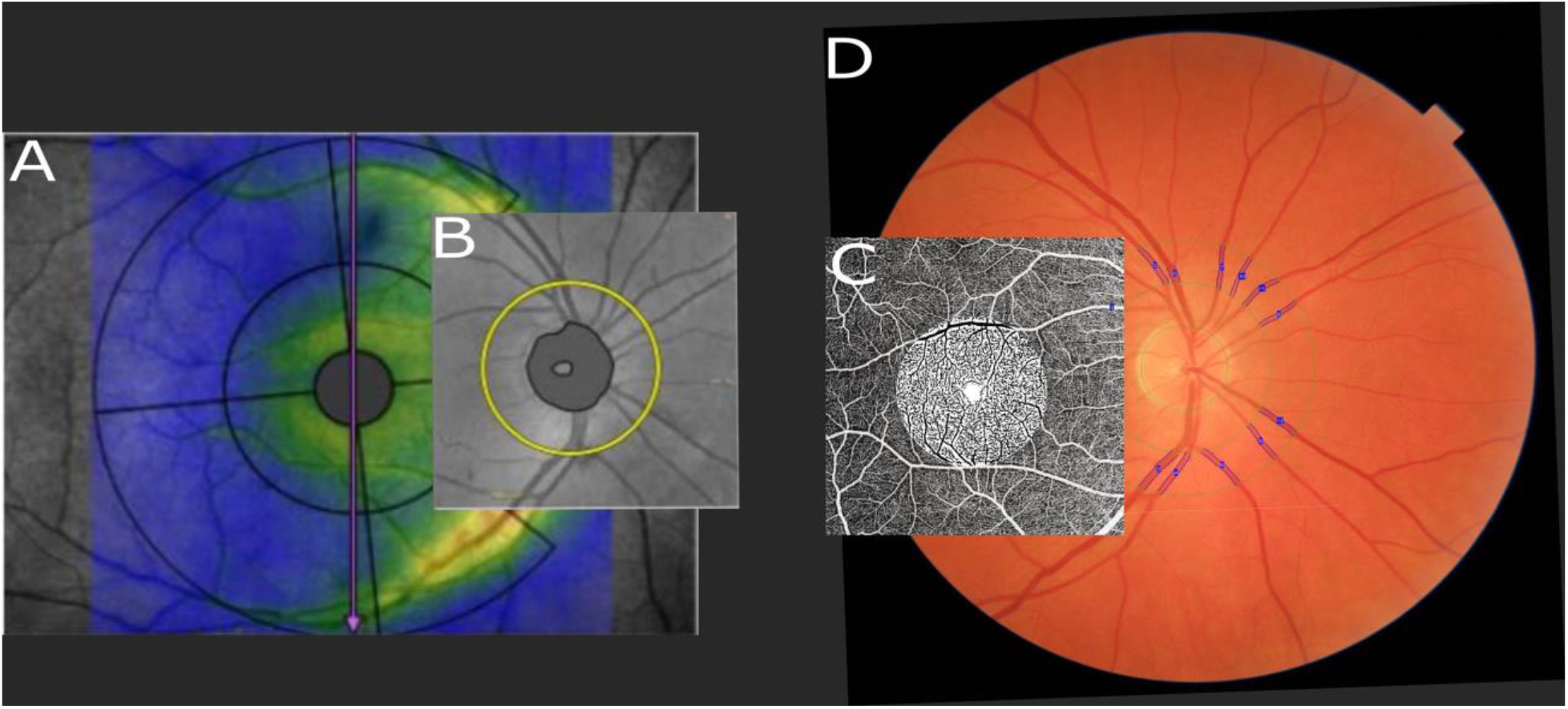
Structural (A-B) and vascular (C-D) regions of interest. A) Macular retinal nerve fiber layer (mRNFL) and ganglion cell-inner plexiform layer (GCIPL) thicknesses measured within the larger (10-mm diameter) circle centered at the fovea, excluding the innermost 1-mm diameter circle and the nasal sector of the outer ring. B) Peripapillary RNFL (pRNFL) measured at a circle of 3.45-mm diameter centered at the optic nerve head (ONH). C) Optical Coherence Tomography-Angiography (OCT-A) scan of 6×6 mm centered at the fovea. Signal intensity inside the innermost 3-mm diameter circle is binarized in flow (black) and non-flow (white). D) 45° fundus image centered at the ONH. The 6 largest arterioles and 6 largest venules between rings of 2 and 3 optic disc diameters are marked in blue.

After registering and binarizing the signal of the en face OCT-A images, we calculated the fractal dimension (FD) of the superficial vascular plexus, inside a 3-mm diameter circle centered at the fovea (Figure 1C). We have previously provided details on FD and its calculation, as well as on the specifications and repeatability of the Canon OCT-A.^28,38^ In short, FD represents the complexity of the branching pattern and is lower in conditions with sparser vasculature, such as glaucoma.^39^

Lastly, we acquired two 45° high-quality and artifact-free fundus images (TRC-NW400, Topcon Corporation, Tokyo, Japan), centered at the ONH (Figure 1D). For each image, we derived the central retinal artery and vein equivalents (CRAE, CRVE; i.e., diameters) using the standardized Knudtson-Parr-Hubbard iteration, whose details and validation can be found elsewhere.^28,40^ In short, we back-calculated vessel diameters using the 6 largest arteriolar and 6 largest venular branches, identified within a ring centered at the ONH (2 and 3 optic disc diameters). We recorded the average CRAE and CRVE of two images.

### Retinal blood flow and lower autoregulation limit

We calculated total retinal vascular resistance (RVR) using the measured FD, CRAE, and CRVE of each participant, as well as population-based hematocrit values (Lifelines Biobank), adjusted for age, sex, and blood pressure status. We have previously documented the mathematics behind this Poiseuille-based model and its validation in vivo.^28,41^ Subsequently, we computed total RBF, using RVR and retinal perfusion pressure (RPP), a more precise estimation of OPP for the retinal circulation^28^:

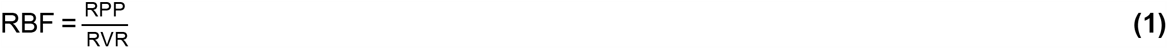

where RPP = (0.39·MAP + 10.1) - IOP mmHg and 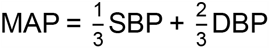 is the mean arterial pressure.

We defined LARL as the lowest RPP value for which RBF can be maintained constant (Figure 2). At this critical point, the vasculature has reached its maximal autoregulatory capacity and any further pressure drop will not trigger compensatory vasodilation, resulting in flow reduction. We have previously shown that LARL can be approximated as:

**Figure 2.**
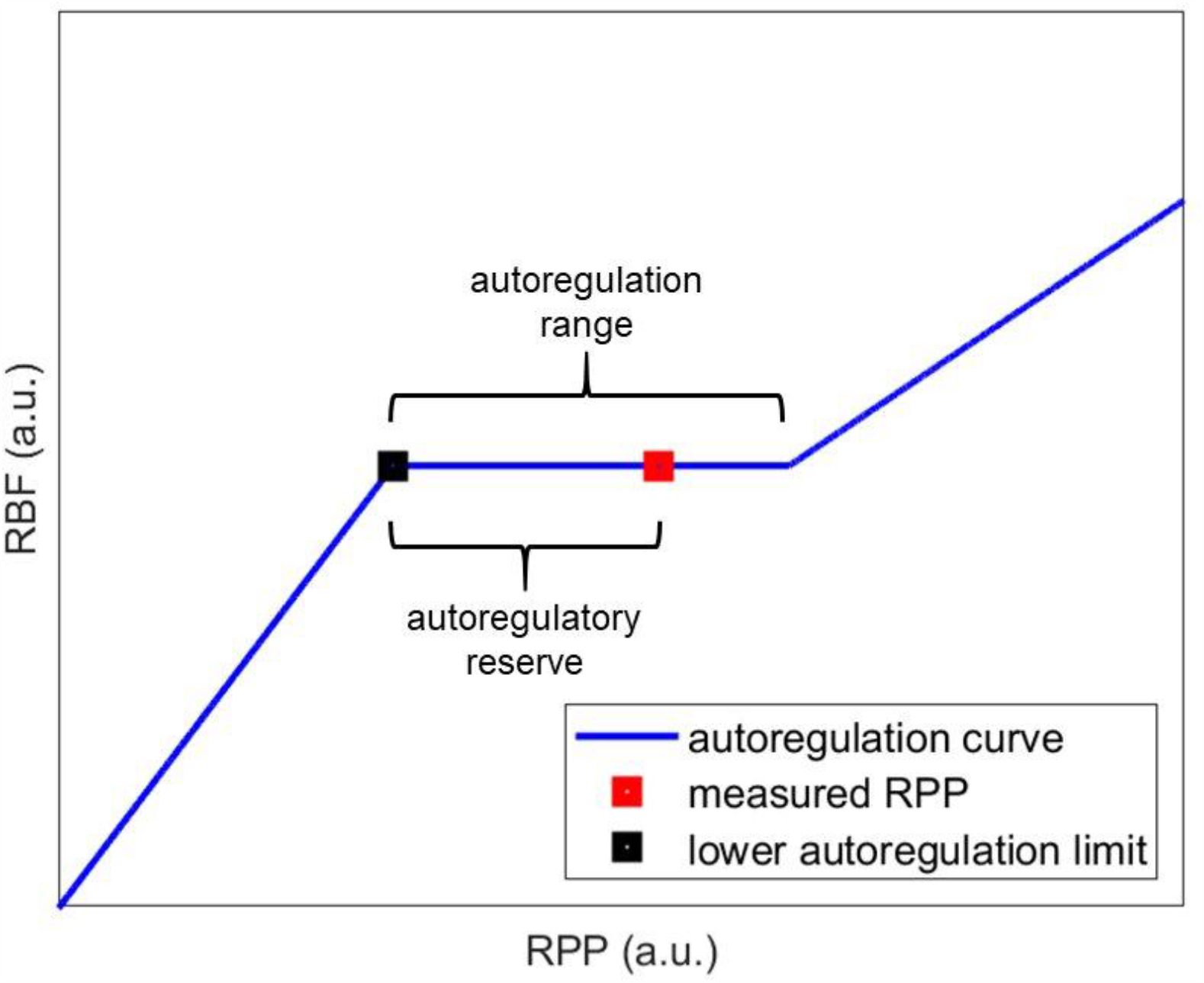
Theoretical autoregulation curve (axes in arbitrary units). Retinal blood flow (RBF) is displayed as a function of retinal perfusion pressure (RPP). Within the autoregulation range, blood flow is maintained constant. The distance of the actual (measured) RPP from the lower autoregulation limit (LARL) is the autoregulatory reserve (AR).

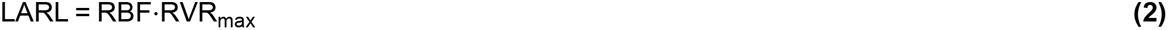

where RVR_max_ is an upper bound observed in a population.^28^

In this study, we defined RVR_max_ as the 95th percentile of the RVR distribution. Due to the possible occurrence of structural remodeling in retinal vessels belonging to subjects with AHT, we separated the RVR distributions of the non-hypertensives (groups 1 and 2) and hypertensives (groups 3 and 4).^12^

Lastly, for each participant, we defined the *autoregulatory reserve* (AR) as the difference between measured RPP and predicted LARL (Figure 2).

### Statistical analysis

For the first part of the analysis, to establish the existence of a U-shaped association (if any), we univariably compared structural OCT metrics (mRNFL, GCIPL, and pRNFL) between the four BP groups. For the second part of the analysis, to investigate whether any vascular factors could possibly explain this association, we univariably compared RVR, RBF, and AR between the groups. For the last part of the analysis, we performed mediation analysis to examine whether the vascular metrics lie in the explanatory pathway of the relationship between MAP and the structural OCT metrics that were significant in the first part of the analysis.

We described normally distributed variables with the mean and standard deviation (SD) and variables with a skewed distribution with the median and interquartile range (IQR). We used one-way ANOVA with post hoc tests for group mean comparisons, adjusting for potential confounders. To account for multiple testing, we implemented the Tukey HSD correction. We applied Levene’s test to check for equality of variances. Whenever ANOVA assumptions were not met, we used non-parametric tests, Welch’s one-way ANOVA with the Games-Howell correction, or quantile regression.

To determine a mediation effect, we used Baron and Kenny’s mediation steps.^42^ In short, a vascular factor M was considered a mediator of the effect of MAP (X) on structural OCT (Y), if the following were true in linear regression analysis:

a. X was a significant predictor of Y (Y∼X).
b. X was a significant predictor of M (M∼X).
c. When M was added to the model (Y∼X+M), M was a significant predictor of Y and the significance of X as a predictor of Y was reduced.

We verified these findings by using the Sobel test for indirect effects. Mediation analysis was performed separately for the low BP group together with the controls, the treated AHT group together with the controls, and the untreated AHT group together with the controls. RBF, RVR, and AR were examined as potential mediators.

All analyses were performed using R (version 3.3.3; R Foundation for Statistical Computing, Vienna, Austria) and SPSS (version 26; IBM Corp., Armonk, NY). A *P* value of 0.05 or less was considered statistically significant.

## Results

In total, 96 eyes of 96 subjects fulfilled all the criteria and were included in the analysis. Table 1 displays the characteristics of the population, stratified by BP status. Sex and BMI were significantly different between groups, aside from BP. As expected, the low BP group comprised almost exclusively females, while higher BMI was present in the hypertensive groups.^43,44^ Other factors that could affect the comparisons, such as age, IOP, spherical equivalent (SEQ), and ONH area, were similar between groups.

**Table 1.** Characteristics of the study population.

|  | **Group 1**<br>**(Low BP)** | **Group 2**<br>**(Normal BP)** | **Group 3**<br>**(Treated AHT)** | **Group 4**<br>**(Untreated AHT)** | ***P* value** |
| --- | --- | --- | --- | --- | --- |
| **Group size (*N*)** | 31 | 21 | 26 | 18 |  |
| **Age; years**<br>**[mean (SD)]** | 56.1 (4.4) | 55.9 (4.7) | 56.4 (4.8) | 57.2 (4.6) | 0.81 |
| **Sex; % female** | 93.5 | 47.6 | 42.3 | 44.4 | **<0.001** |
| **SBP; mmHg**<br>**[mean (SD)]** | 106 (9) | 126 (6) | 142 (18) | 159 (22) | **<0.001** |
| **DBP; mmHg**<br>**[mean (SD)]** | 66 (6) | 79 (6) | 86 (11) | 99 (8) | **<0.001** |
| **BMI; kg·m<sup>-2</sup>**<br>**[median (IQR)]** | 22.1<br>(21.2 to 24.3) | 23.3<br>(22.1 to 26.5) | 26.9<br>(24.7 to 29.8) | 27.3<br>(24.3 to 28.4) | **<0.001** |
| **Smoking; % ever** | 22.6 | 38.1 | 30.8 | 38.9 | 0.57 |
| **IOP; mmHg**<br>**[mean (SD)]** | 13.9 (3.0) | 13.2 (3.1) | 14.3 (3.0) | 14.6 (3.7) | 0.56 |
| **SEQ; D**<br>**[mean (SD)]** | -0.10 (1.41) | +0.27 (1.67) | -0.23 (1.55) | -0.68 (1.69) | 0.31 |
| **ONH area; mm<sup>2</sup>**<br>**[median (IQR)]** | 1.89<br>(1.69 to 2.24) | 1.96<br>(1.71 to 2.20) | 1.94<br>(1.72 to 2.31) | 2.00<br>(1.78 to 2.29) | 0.75 |
| BP, blood pressure; AHT, arterial hypertension; SD, standard deviation; SBP, systolic blood pressure; DBP, diastolic blood pressure; BMI, body mass index; IOP, intraocular pressure; SEQ, spherical equivalent; ONH, optic nerve head. |  |  |  |  |  |

### Structural metrics

Table 2 and Figure 3 present the comparison of structural OCT metrics across the four BP groups. Adjusted post hoc comparisons revealed that, compared to the controls, GCIPL was significantly thinner in the low BP group (*P*_*adj*_ = 0.013), the treated AHT group (*P*_*adj*_ = 0.007), and the untreated AHT group (*P*_*adj*_ = 0.007). The mRNFL was also thinner, but this was only significant for the treated AHT group (*P*_*adj*_ = 0.001). Interestingly, mRNFL in treated hypertensives was even significantly thinner than in untreated hypertensives (*P*_*adj*_ = 0.033). Figure 3 shows the characteristic (inverse) U shape for the macular OCT metrics. There was no clear effect of BP group on the mean pRNFL. However, treated hypertensives had a thinner temporal pRNFL (*P*_*adj*_ = 0.045) than normotensives. Also, inferior pRNFL was borderline thinner in subjects with low BP (*P*_*adj*_ = 0.083) and clearly thinner in both treated and untreated hypertensives (*P*_*adj*_ = 0.034 and 0.033, respectively). Sex and BMI did not confound any of the associations (all *P* values >> 0.05) and this was still true after the omission of any group from the analysis.

**Table 2.**
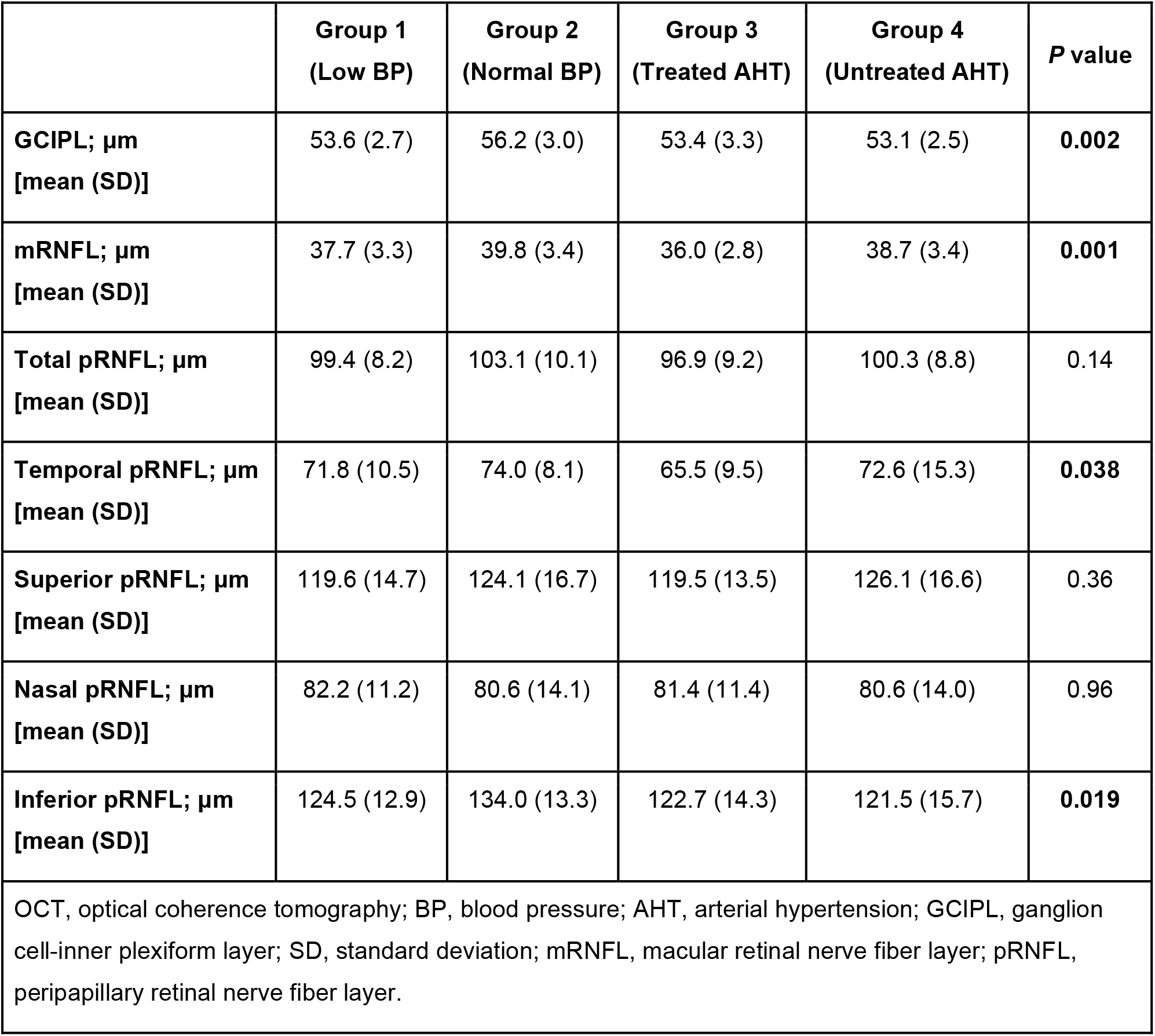
Structural OCT metrics as a function of BP status.

**Figure 3.**
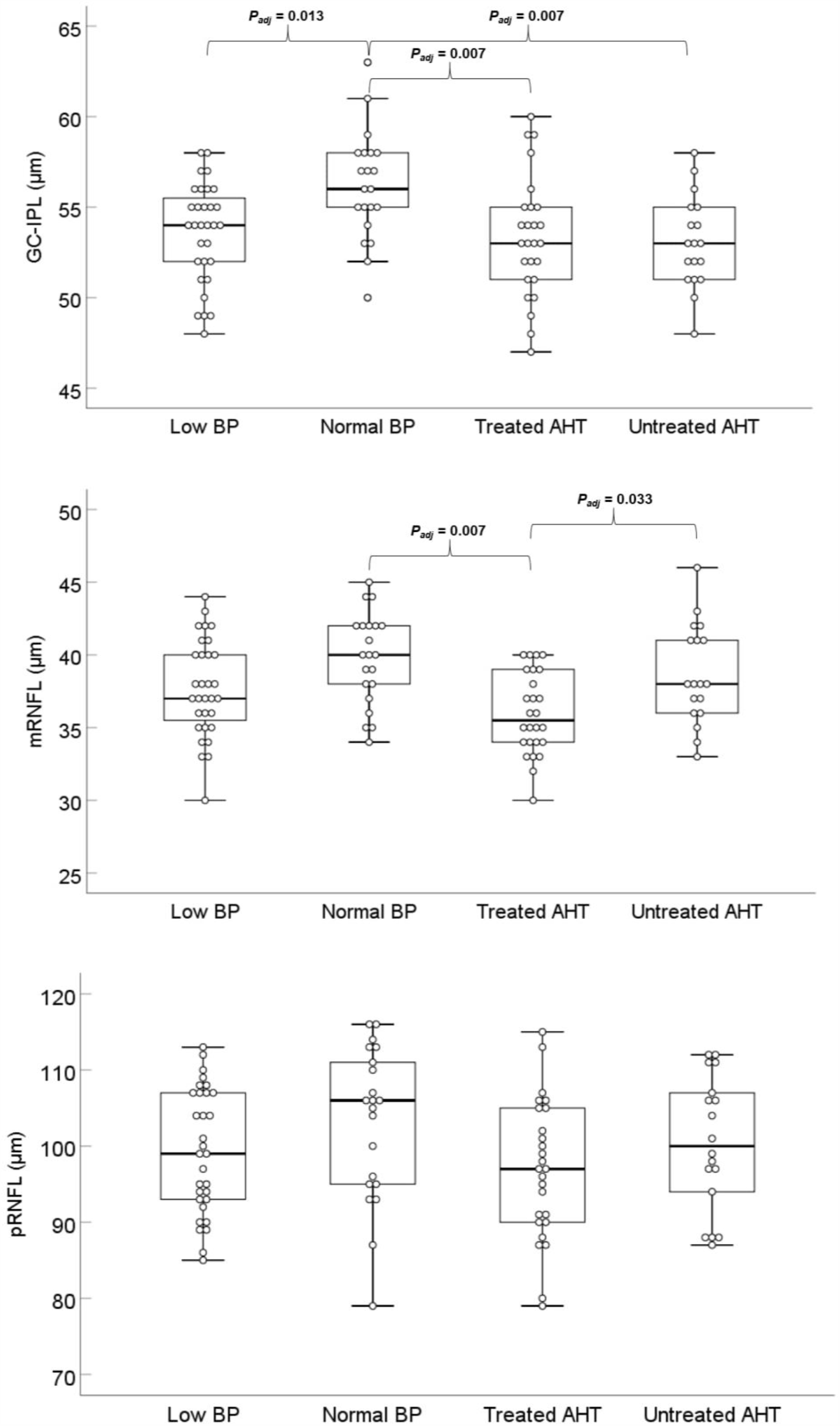
Ganglion cell-inner plexiform layer (GCIPL), macular retinal nerve fiber layer (mRNFL), and peripapillary retinal nerve fiber layer (pRNFL) as a function of blood pressure (BP) status. Significant differences after post hoc comparisons (adjusted for multiple testing) are marked. The thicker layers observed in the control group (normal BP) than in the low BP or arterial hypertension (AHT) groups create a characteristic (inverse) U shape.

### Vascular metrics

Figure 4 displays RBF, RVR, and AR as a function of BP status. There were differences in RBF between groups (*P* = 0.034), but after adjusting for multiple comparisons RBF was only significantly lower in the low BP group when compared to the untreated AHT group (*P*_*adj*_ = 0.043). RVR was also different between groups (*P* = 9.0·10^−9^), with the additional presence of a larger variance in the treated AHT group (Levene’s test: *P* = 0.002). With regards to AR, the unequal variances were also statistically significant (Levene’s test: *P* = 0.0002), showing that, unlike any other group, treated hypertensives could have either a large or a small AR. As can be better seen in Figure 5 and Supplementary Table S1, the low BP group had a significantly smaller AR than the control group, regardless of AR quantile compared. Conversely, the untreated hypertensives had a significantly larger AR than the control group, regardless of quantile compared. However, there was a mixed response in the treated AHT group: the AR was significantly smaller than that of the controls for small quantiles, while it was similar or larger for larger quantiles. In addition, correlation analysis within the treated AHT group revealed that the smaller AR quantiles corresponded to the lowest MAP values (Pearson’s r = 0.45, *P* = 0.020), i.e., to the most intensively controlled hypertensives. Again, sex and BMI did not confound these associations.

**Figure 4.**
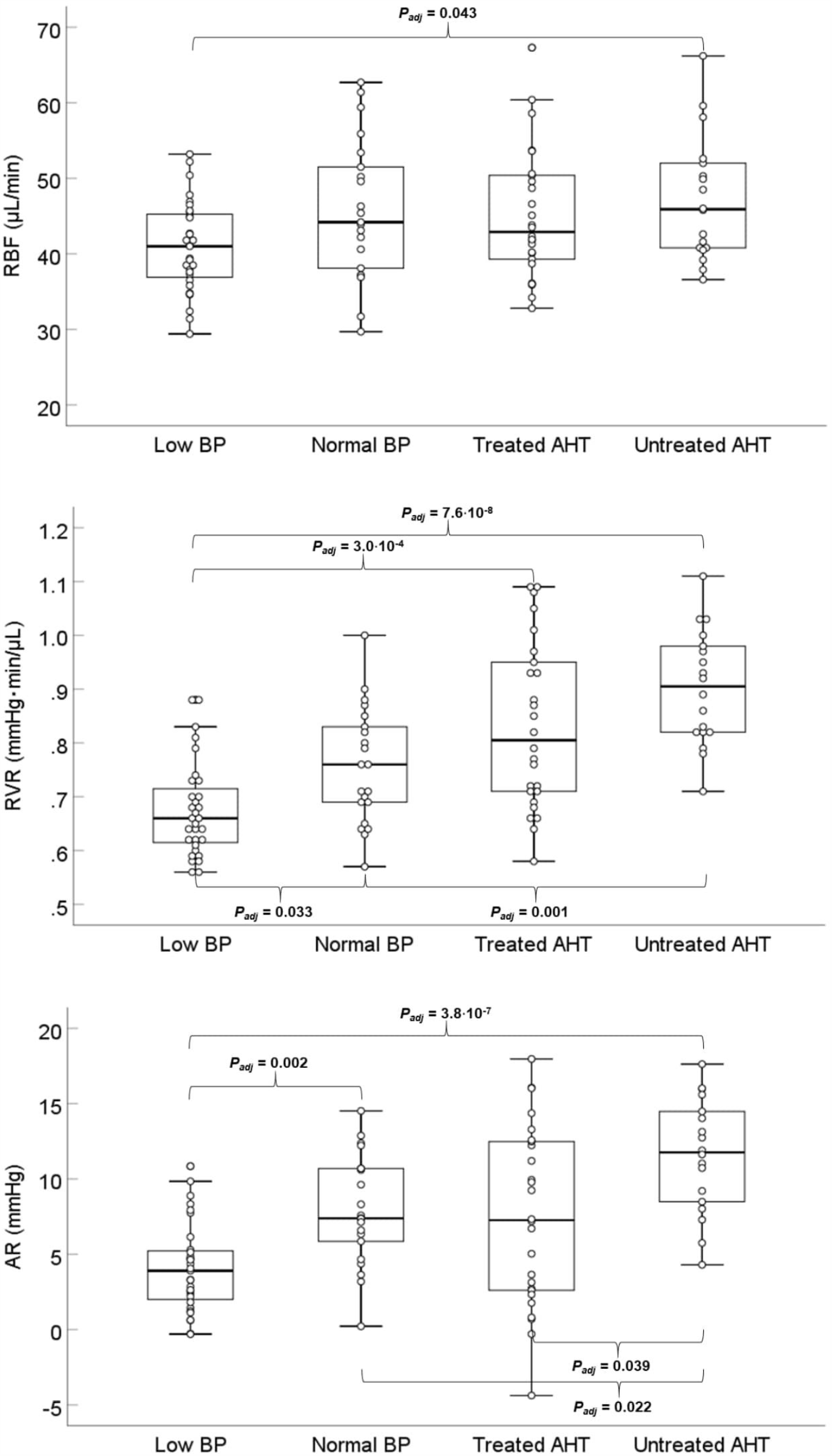
Absolute total retinal blood flow (RBF), retinal vascular resistance (RVR), and autoregulatory reserve (AR) as a function of blood pressure (BP) status. Significant differences after post hoc comparisons (adjusted for multiple testing) are marked. With increasing BP, RBF increases less than RVR, resulting in an autoregulation effect visible in the first panel. A statistically significant larger variability is observed for the AR of treated hypertensives (third panel), suggesting that subjects in this group can be very close or very far from the lower autoregulation limit.

**Figure 5.**
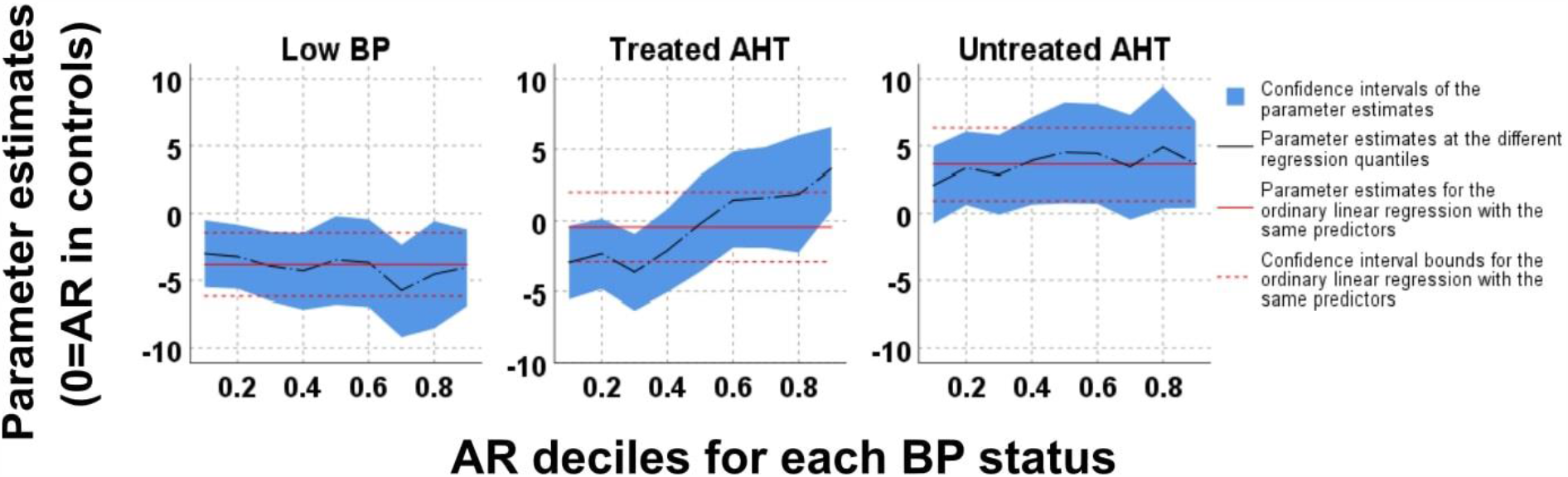
Quantile regression models for autoregulatory reserve (AR) at each blood pressure (BP) status. Parameter estimates (y-axis) represent the relative AR (compared to controls) for every AR decile (x-axis). Low BP individuals (left panel) have a smaller AR than controls (y<0), while individuals with untreated arterial hypertension (AHT; right panel) have a larger AR than controls (y>0). Individuals with treated AHT (middle panel) have a smaller AR only when intensively treated (leftmost deciles).

### Mediation analysis

Results from mediation analysis regarding the effect of BP status on GCIPL are presented in Table 3. RBF was mediating the association of GCIPL with BP within the combined low BP group and controls, while RVR was mediating the same association within the combined untreated AHT group and controls. RVR and AR were both independently mediating the association of GCIPL with BP within the combined treated AHT group and controls. In the complete model (GCIPL∼MAP+RVR+AR), which accounts for the covariance between RVR and AR, the opposite, real effect of AR became visible, that is, small AR was associated with thinner GCIPL (see Discussion section). We did not observe any vascular mediation for the effect of BP status on RNFL metrics.

**Table 3.**
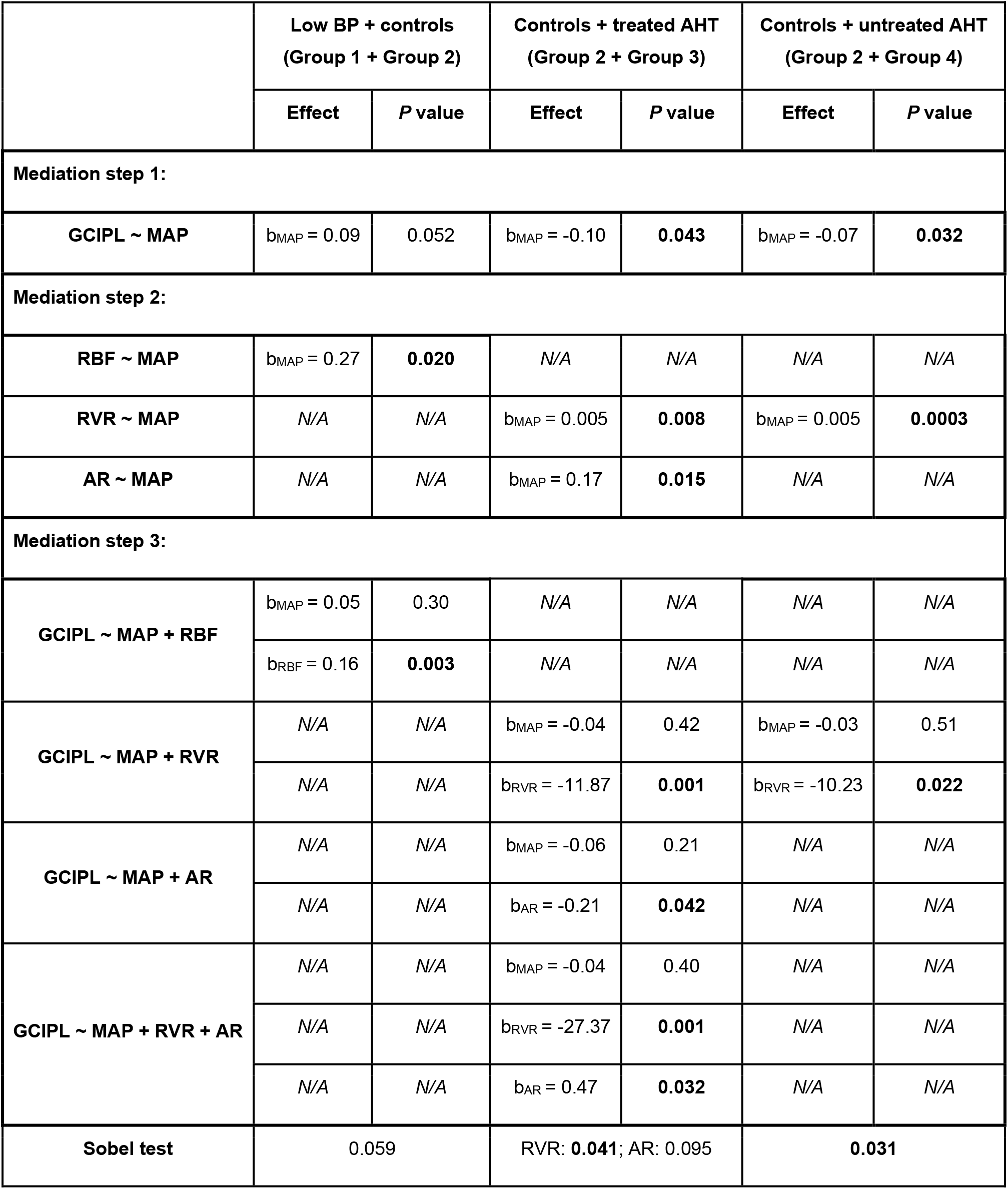
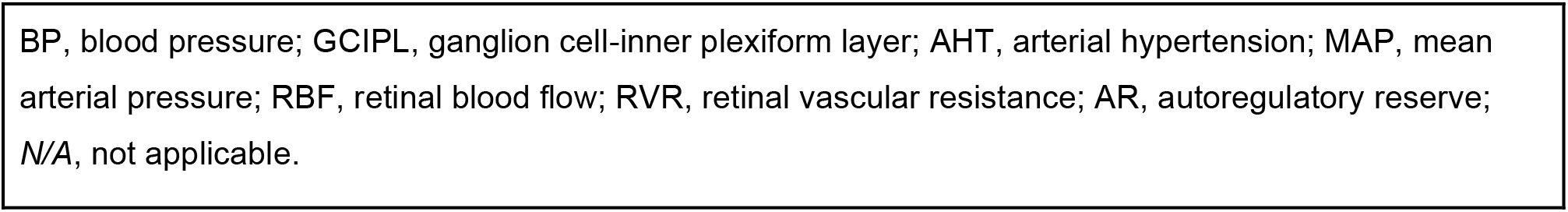
Effect of BP status on GCIPL: mediation analysis.

|  | Low BP + controls<br>(Group 1 + Group 2) |  | Controls + treated AHT<br>(Group 2 + Group 3) |  | Controls + untreated AHT<br>(Group 2 + Group 4) |  |
| --- | --- | --- | --- | --- | --- | --- |
|  | Effect | P value | Effect | P value | Effect | P value |
| **Mediation step 1:** |  |  |  |  |  |  |
| **GCIPL ~ MAP** | $b_{MAP} = 0.09$ | 0.052 | $b_{MAP} = -0.10$ | **0.043** | $b_{MAP} = -0.07$ | **0.032** |
| **Mediation step 2:** |  |  |  |  |  |  |
| **RBF ~ MAP** | $b_{MAP} = 0.27$ | **0.020** | N/A | N/A | N/A | N/A |
| **RVR ~ MAP** | N/A | N/A | $b_{MAP} = 0.005$ | **0.008** | $b_{MAP} = 0.005$ | **0.0003** |
| **AR ~ MAP** | N/A | N/A | $b_{MAP} = 0.17$ | **0.015** | N/A | N/A |
| **Mediation step 3:** |  |  |  |  |  |  |
| **GCIPL ~ MAP + RBF** | $b_{MAP} = 0.05$ | 0.30 | N/A | N/A | N/A | N/A |
| | $b_{RBF} = 0.16$ | **0.003** | N/A | N/A | N/A | N/A |
| **GCIPL ~ MAP + RVR** | N/A | N/A | $b_{MAP} = -0.04$ | 0.42 | $b_{MAP} = -0.03$ | 0.51 |
| | N/A | N/A | $b_{RVR} = -11.87$ | **0.001** | $b_{RVR} = -10.23$ | **0.022** |
| **GCIPL ~ MAP + AR** | N/A | N/A | $b_{MAP} = -0.06$ | 0.21 | N/A | N/A |
| | N/A | N/A | $b_{AR} = -0.21$ | **0.042** | N/A | N/A |
| **GCIPL ~ MAP + RVR + AR** | N/A | N/A | $b_{MAP} = -0.04$ | 0.40 | N/A | N/A |
| | N/A | N/A | $b_{RVR} = -27.37$ | **0.001** | N/A | N/A |
| | N/A | N/A | $b_{AR} = 0.47$ | **0.032** | N/A | N/A |
| **Sobel test** | 0.059 |  | RVR: **0.041**; AR: 0.095 |  | **0.031** |  |

BP, blood pressure; GCIPL, ganglion cell-inner plexiform layer; AHT, arterial hypertension; MAP, mean arterial pressure; RBF, retinal blood flow; RVR, retinal vascular resistance; AR, autoregulatory reserve; N/A, not applicable.

## Discussion

In this study, we reported three main findings. Firstly, there exists an inverse U-shaped effect between blood pressure status and structural OCT metrics (GCIPL and RNFL), with both low and high blood pressure being associated with thinning of the inner retinal layers. Secondly, despite the existence of retinal blood flow autoregulation, only a small autoregulatory reserve is present in individuals with low blood pressure, as well as in individuals with intensively treated arterial hypertension. Lastly, this compromised capacity for retinal blood flow regulation explains (mediates) the effect of blood pressure status on the GCIPL.

### Low blood pressure

This is, to our knowledge, the first study to uncover an association between low BP and thinning of the inner retina in ophthalmologically healthy subjects. This relationship and its vascular mediation were more pronounced for the GCIPL, which has been shown to be the main layer of early NTG manifestation.^45^ In addition, the association was entirely mediated by RBF (no effect of sex, BMI, or other confounders in our population).

Indeed, we have previously shown that LARL for subjects without AHT corresponds to a realistic SBP/DBP of ∼105/65 mmHg (or even higher if IOP is above average).^28^ Since, in the present study, the average BP reading for the low BP group was at 106/66 mmHg, our finding that this group had a borderline lower RBF (Figure 4, Table 3) and a considerably smaller AR (Figure 5, Table S1) than controls is in line with our estimations and the general concept of autoregulation.

Population studies have failed to report this association between low BP and GCIPL thickness, possibly due to the implementation of linear models, but an explanation due to differences in genetic background cannot be excluded.^33,34^ However, nonlinear models were used is studies with glaucoma as the outcome measure and, in line with our findings, there is evidence for the existence of increased glaucoma risk with low, usually diastolic or nocturnal, BP. ^13–16,18,46^

### Hypertension

Despite a slightly higher RBF and a considerably higher AR (Figures 4 and 5), untreated hypertensives had thinner GCIPL, which has also been shown to be the location of first progression in glaucoma patients with AHT.^45^ This was mediated by RVR (Table 3), i.e., the negative effect of increased BP to the GCIPL is explained by increased vascular resistance (but not by reduced blood flow - see below). Nevertheless, it was the treated AHT group that exhibited the most pronounced thinning and this was present in the majority of structural OCT metrics (GCIPL, mRNFL, temporal pRNFL, inferior pRNFL). For this group, GCIPL thinning was independently mediated by RVR and AR (Figure 5, Table 3). In the univariable models, large RVR and large AR were both associated with thinner GCIPL, due to substantial covariance. After controlling for the confounding effect of RVR, small AR was associated with thinner GCIPL. This suggests that, in treated AHT, a combination of increased resistance and being close to the autoregulatory tipping point explain the negative effect to the GCIPL.

GCIPL thinning without a decrease in RBF seems counterintuitive. However, even when total RBF is largely unaffected, increased RVR results in increased blood velocity (i.e., reduced transit time), shunting of flow, and reduced capillary density.^47,48^ This could affect red blood cell distribution and retinal oxygen extraction. Smaller AR was additionally present in intensively treated subjects (Figure 4C, Figure 5, Table S1), which could mimic low BP and lead to hypoperfusion of the RGCs. In this regard, our results reflect a possible effect of the combined rightward shift of the autoregulation curve (due to atherosclerosis and arteriosclerosis) and the variations of the measured RPP (Figure 2), due to BP fluctuations throughout the day. Lastly, chronic AHT also results in endothelial dysfunction and, therefore, impaired autoregulation.^49^

A number of previous studies have also shown an effect of AHT on GCIPL and RNFL thickness.^21,29–32^ However, some population-based studies did not detect this relationship.^33,34^ Again, study design and analysis methods seem to be the most likely explanations for this discrepancy. In all, the existence of thinner GCIPL in both low BP and AHT creates the characteristic inverse U-shaped association. Regarding glaucoma risk, the role of AHT is controversial. Most evidence points towards at least some benefit of timely AHT treatment, possibly due to the prevention of microvascular damage, in combination with a slight IOP lowering.^24,50^ However, it has been suggested that aggressive treatment of AHT could negatively affect glaucoma and our results also point towards this direction.^20^ The confounding contribution of individual antihypertensive medications, whether neuroprotective or detrimental, remains inconclusive.^21–25^

We would like to stress here that, since there are benefits to intensive BP control with regards to cardiovascular disease, our results should not be considered as a case for milder treatment of AHT in general.^51,52^ However, since a J or U-shaped effect is reported in both fields when intensive treatment becomes too intensive, our results could be a starting point for discussion with cardiologists in individual cases where, for example, glaucoma continues to deteriorate despite adequate IOP control.^53^

### ‘Chicken-egg’ dilemma

From a theoretical standpoint, there exists a point in the pre-disease time course when the very first vascular deficits or the very first structural deficits manifest. A subsequent causal cascade of events would then result in further mutually mediated vascular and structural deterioration, sometimes leading to a glaucoma diagnosis. By definition, cross-sectional studies regarding perfusion in glaucoma cases are well past this critical point and cannot enhance our understanding of the process.

In this regard, our study’s novelty lies in demonstrating that interdependent structural and vascular deficits related to a long-debated cardiovascular risk factor (especially low BP) can even be traced back to whom we perceive as ophthalmologically healthy subjects. These results suggest that a subtle GON precursor remains rather elusive, albeit present early on. While this observation adequately explains the claim that perfusion deficits are already present prior to glaucoma, it does not fully resolve the ‘chicken-egg’ dilemma (we showed that vascular deficits are present without glaucoma, but not necessarily without smaller structural deficits). However, the existence of different mediators between MAP and GCIPL thickness with different BP status, hints towards a preceding vascular component. For example, there is no parsimonious explanation for an intensive antihypertensive treatment strategy following GCIPL thinning.

### Study strengths and limitations

The main strength of this study was the strict selection process which allowed us to look at the true extremes of BP. This reduces the noise that usually characterizes larger population studies and results in indirect loss of power. In addition, our linearity-free assumptions and the categorizing of BP (rather than considering it as a continuous variable) allowed us to differentiate between BP status and uncover a U-shaped association that was previously elusive. Lastly, to our knowledge, this study is the first to provide a rigorous explanation of the differential effect of BP status on retinal structure, by directly linking it to total RBF and its autoregulation.

Due to the cross-sectional nature of our study, absence of data on the first occurrence of AHT is a limitation. However, our threshold of an at least 1-year old diagnosis, together with the selection procedure using multiple previous visits from another database, ensured no newly-diagnosed cases (almost all cases had been diagnosed before at least 3 years). Secondly, RBF, RVR, and AR were not directly measured, but indirectly calculated by means of their measured components and a previously validated model. Indeed, we have previously shown in an independent validation dataset that these outcomes strongly correlate with in vivo blood flow metrics, as well as with structural metrics.^28^ Unfortunately, there is currently no gold standard way to quantify these parameters, but Doppler OCT is a frequently-used, promising tool. Our predictive approach has the advantage of using more reproducible imaging techniques and, most importantly, taking into account autoregulation limits. Combining these two methods could, therefore, further finetune estimations. It should be also noted that our approach provides information on the effect of static RBF autoregulation, but it is possible that BP status also results in impairment of the autoregulatory latency, i.e., dynamic autoregulation. As such, our results might only be part of a bigger underlying effect. Lastly, our population was predominantly Caucasian; it is to be determined if the results can be generalized to other ethnicities.

In conclusion, we uncovered thinning of the GCIPL and RNFL related to both tails of the blood pressure distribution (inverse U-shaped effect) and to intensive treatment of AHT, in ophthalmically healthy individuals. We found that GCIPL thinning was associated with reduced autoregulatory capacity of retinal blood flow. This predisposition to glaucomatous damage could explain the frequent epidemiological finding of increased glaucoma risk in certain population subgroups, such as subjects with nocturnal blood pressure dipping or aggressively treated hypertension. Longitudinal studies are needed to confirm this postulation.

## Data Availability

Data is freely available upon request to the corresponding author (Konstantinos Pappelis).

## Abbreviations/Acronyms

AHT: arterial hypertension;
AR: autoregulatory reserve;
BMI: body mass index;
BP: blood pressure;
CRAE: central retinal artery equivalent;
CRVE: central retinal vein equivalent;
DBP: diastolic blood pressure;
FD: fractal dimension;
GCIPL: ganglion cell-inner plexiform layer;
IOP: intraocular pressure;
IQR: interquartile range;
LARL: lower autoregulation limit;
MAP: mean arterial pressure;
mRNFL: macular retinal nerve fiber layer;
NTG: normal-tension glaucoma;
OCT: optical coherence tomography;
OCT-A: optical coherence tomography-angiography;
ONH: optic nerve head;
OPP: ocular perfusion pressure;
pRNFL: peripapillary retinal nerve fiber layer;
RBF: retinal blood flow;
RGC: retinal ganglion cell;
RNFL: retinal nerve fiber layer;
RPP: retinal perfusion pressure;
RVR: retinal vascular resistance;
SBP: systolic blood pressure;
SD: standard deviation;
SEQ: spherical equivalent.

