## Supplementary Table S1 for "U-shaped effect of blood pressure on structural OCT metrics and retinal blood flow autoregulation in ophthalmologically healthy subjects"

**Supplementary Table S1.** AR as a function of BP status, compared to controls per decile.

|  | **Low BP (Group 1)**  *Baseline: Group 2* | | **Treated AHT (Group 3)**  *Baseline: Group 2* | | **Untreated AHT (Group 4)**  *Baseline: Group 2* | |
| --- | --- | --- | --- | --- | --- | --- |
|  | **Effect (b)** | ***P* value** | **Effect (b)** | ***P* value** | **Effect (b)** | ***P* value** |
| Q10 | -3.0 | **0.019** | -3.0 | **0.027** | 2.1 | 0.15 |
| Q20 | -3.2 | **0.009** | -2.3 | 0.065 | 3.3 | **0.026** |
| Q30 | -3.9 | **0.003** | -3.6 | **0.009** | 2.9 | 0.057 |
| Q40 | -4.3 | **0.004** | -2.1 | 0.16 | 3.9 | **0.019** |
| **Q50** | -3.5 | **0.038** | -0.2 | 0.91 | 4.5 | **0.019** |
| Q60 | -3.7 | **0.027** | 1.5 | 0.39 | 4.4 | **0.020** |
| Q70 | -5.8 | **0.001** | 1.6 | 0.36 | 3.4 | 0.083 |
| Q80 | -4.6 | **0.025** | 1.9 | 0.37 | 4.9 | **0.034** |
| Q90 | -4.0 | **0.006** | 3.6 | **0.016** | 3.6 | **0.027** |
| AR, autoregulatory reserve; BP, blood pressure; AHT, arterial hypertension. | | | | | | |
